# Severe Acute Respiratory Syndrome Coronavirus 2 Serology in Asymptomatic Healthcare Professionals: Preliminary Experience of a Tertiary Italian Academic Center

**DOI:** 10.1101/2020.04.27.20073858

**Authors:** F Tosato, M Pelloso, N Gallo, C Giraudo, G Llanaj, C Cosma, C Pozzato, A Padoan, D Donato, M Plebani

## Abstract

We investigated the SARS-CoV-2 specific antibody titers in 133 asymptomatic healthcare providers working at the Department of Laboratory Medicine of our tertiary center. A commercial chemiluminescence immunoassay, validated according to the ISO15189 standard requirements, was used. All the enrolled healthcare professionals underwent, simultaneously to the blood sampling, a nasopharyngeal swab for molecular testing with quantitative reverse-transcriptase-based polymerase chain reaction (RT-PCR). An overall positiveness of 5.25% was found. We strongly promote a wide use of validated serologic assays in asymptomatic, healthy individuals, as a crucial information for epidemiological surveillance.

---

The current pandemic of coronavirus disease 2019 (COVID-19) requires a global and tremendous effort not only to treat the symptomatic patients but also to implement active surveillance programs and assure the development and use of accurate diagnostic, prognostic, and therapeutic tools^1^.

Considering the preliminary published evidence and the well-known role of serological analysis especially for serosurveillance studies^2,3^, we investigated the severe acute respiratory syndrome coronavirus 2 (SARS-CoV-2) specific antibody titers in 133 healthcare providers working at the Department of Laboratory Medicine of our tertiary center which is characterized by a workload of around 34,000 tests per day of both inpatients and outpatients.

It has to be underlined that in accordance with the recommendations of the World Health Organization, personal protective equipment together with social distancing and preventive hygiene measures were applied by all our staff since the spread of the pandemic in our country.

All the enrolled healthcare professionals underwent, simultaneously to the blood sampling, a nasopharyngeal swab for molecular testing and the quantitative reverse-transcriptase-based polymerase chain reaction (RT-PCR) was applied.

The study was submitted to the Ethical Committee of the University-Hospital of Padova (protocol number 23307).

IgM and IgG antibodies against SARS-CoV-2 in serum samples were tested using the MAGLUMI 2019-nCoV IgM/IgG kit, an automated central laboratory rapid test that runs on the MAGLUMI chemiluminescence immunoassay system, which received the CE mark on the 19th of February 2020 and is supplied by Snibe Diagnostic (Shenzhen, China). The validation of the method was performed according to the ISO15189 standard requirements. The quality of conformance declared by the manufacturer regarding serum inactivation, intra- and inter-day repeatability, and linearity was confirmed^4,5^. Diagnostic sensitivity and specificity provided by the manufacturer and deriving from two Chinese clinical studies are as follows: clinical sensitivity and specificity 78.65% and 97.50% for IgM, 91.21% and 97.33% for IgG, 89.89–95.6% and 96.5–96.0% for IgM+IgG, respectively.

Participants characteristics (sex, age, role and laboratory section) are reported in Table 1.

**Table 1.** Participants characteristics: sex, age, role and laboratory section.

|  | **Total** | **Female** | **Male** |
| --- | --- | --- | --- |
| **Number** | 133 | 117 | 16 |
| **Age, mean (SD)** | 47 (10) | 47 (10) | 47 (13) |
| **Age, median (IQR)** | 51 (39, 55) | 51 (39, 55) | 53 (36, 57) |
| **Role, n (%)** |  |  |  |
| Clinical Pathologist | 31 (23) | 26 (22) | 5 (31) |
| Nurse and nursing assistant | 28 (21) | 27 (23) | 1 (6) |
| Technician | 59 (44) | 50 (43) | 9 (56) |
| Administrative | 15 (11) | 14 (12) | 1 (6) |
| **Laboratory Section, n (%)** |  |  |  |
| Corelab | 72 (54) | 65 (56) | 7 (44) |
| Emergency | 14 (11) | 10 (9) | 4 (25) |
| Clinical Molecular Biology | 10 (8) | 8 (7) | 2 (13) |
| Phlebotomy Center | 37 (28) | 34 (29) | 3 (19) |

Among the examined asymptomatic healthcare professionals working in our unit, only one (0.75%) was positive for SARS-CoV-2 at the RT-PCR. Moreover, all had negative levels of IgM and IgG (negative cut-off <1.000 kAU/L and <1.100 kAU/L, respectively) except six healthcare providers (4.5%) who showed an increased level of IgG (>1.100 kAU/L). The majority of healthcare professionals with positive IgG titers work at the phlebotomy center, as nurses or nursing assistants. All IgM and IgG titers are plotted in Figure 1.

**Figure 1.**
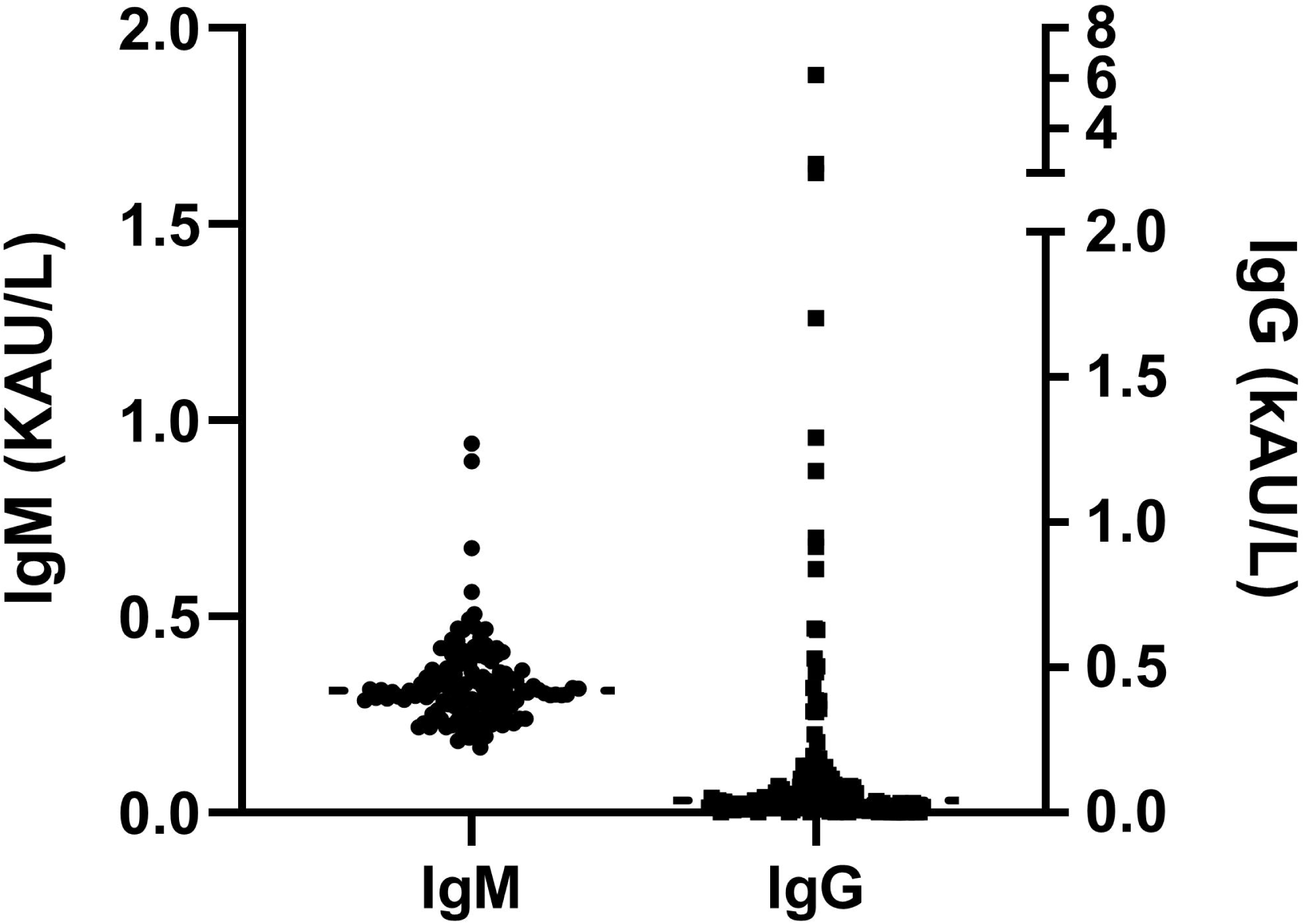

SARS-CoV-2 is a respiratory virus, and as such, it is mainly transmitted between people through respiratory droplets. Therefore, the majority of healthcare professionals with positive IgG titers could easily be expected to work at the phlebotomy center, where there is more contact with patients.

Even if the current knowledge about the antibody response to SAR-CoV-2 is still limited, the published evidence and our preliminary data let us assume that serological analysis may play a significant role. In fact, their contribution could be threefold.

Firstly, the kinetics of immune responses are related to clinical and virological features of patients with COVID-19. Therefore, validated serologic assays can be instrumental for detection of SARSCoV-2–specific antibodies for diagnostic purposes^6–8^.

Moreover, as demonstrated by preliminary evidence, antibodies against SARS-CoV spike may offer some protection against SARS-CoV-2. This is related with the finding that serum samples from convalescent SARS patients tested for their neutralization capacity against SARS-CoV-2, cross-neutralized SARS-2-S-driven entry. This is crucial for evaluating antiviral and vaccine trials, and better understanding of immune protection pathways^8–10^.

Lastly, as mentioned above, the assessment of antibodies seroprevalence is needed for identifying the viral reservoir and for epidemiological studies^8,10,11^. Regarding especially this last aspect, referring to our preliminary data based on a limited population, if we hypothesize analogous results in the Italian population (60.317.000, https://www.istat.it/it/popolazione-e-famiglie 1st January 2020), it would mean that 3,166,642 (5.25%) citizens could be positive although asymptomatic or paucisymptomatic. Obviously, such a direct projection of data cannot be easily made since, in our analysis, we focused on mainly female adults, working in a healthcare environment and wearing protective gears. However, this evidence, if confirmed, as well as general population prevalence estimates, would have a strong impact on the epidemiological management of the disease.

In conclusion, we strongly promote a wide evaluation of IgG and IgM titers using validated serologic assays both in COVID-19 patients as a companion diagnostic tool, but even more in asymptomatic, healthy individuals, as a crucial information for epidemiological surveillance.

## Data Availability

All data referred to in the manuscript are available.

